# Sleep features and long-term incident neurodegeneration: a polysomnographic study

**DOI:** 10.1101/2023.03.23.23287636

**Authors:** Abubaker Ibrahim, Matteo Cesari, Anna Heidbreder, Michaela Defrancesco, Elisabeth Brandauer, Klaus Seppi, Stefan Kiechl, Birgit Högl, Ambra Stefani

## Abstract

While a growing body of studies suggests a link between sleep disturbances and neurodegenerative diseases’ (NDDs) development, prior studies have been hindered by small sample sizes, short follow-up times and a lack of objective sleep measures.

In this cohort study, patients who underwent polysomnography (PSG) at the Innsbruck Sleep Disorders Unit from January 2004 to December 2007, aged ≥18 years, without NDDs at baseline or within five years post PSG, and with at least five years clinical follow-up were included. The main outcome measure was NDDs diagnosis at least five years after polysomnography, assessed until December 2021.

Of 1454 patients assessed for eligibility, 999 (68.7%) met inclusion criteria (683 (68.3%) men; median age 54.9 (interquartile range, IQR 33.9-62.7) years. Seventy-five patients (7.5%) developed NDDs, 924 (92.5%) remained disease-free after 12.8 (IQR 9.9-14.6) years median follow-up. After adjusting for demographic, sleep, and clinical covariates, each percent decrease in sleep efficiency, N3 sleep, or REM sleep was associated with 1.9%, 6.5%, and 5.2% increased risk of incident NDDs (hazard ratio, HR, 1.019, CI:1.002-1.035; HR 1.065, CI:1.007-1.118; HR 1.052, CI:1.012-1.085,), respectively whereas one percent decrease in night-time wakefulness represented a 2.2% reduced risk (HR 0.978, CI:0.958-0.997). Random forest analysis identified wake, followed by N3 and REM sleep percentages, as the most important feature associated with NDDs development. Additionally, multiple sleep features combination offered more robust discrimination of incident NDDs compared to single sleep stages.

These findings support contribution of sleep architecture changes to NDDs pathogenesis and provide insights into the temporal window during which these changes are detectable, pointing to sleep as early NDDs marker and potential target of neuroprotective strategies.

## Introduction

Neurodegenerative diseases (NDDs) constitute one of the greatest challenges of ageing societies, with an enormous impact on affected individuals and their families as well as on health economy. The most prevalent NDDs are Alzheimer’s disease (AD) and Parkinson’s disease (PD), affecting 43.3 and 6.1 million individuals globally^1, 2^, with estimated global medical and non-medical dementia costs of 1.3 US$ trillion in 2019^3^.

Sleep disorders such as insomnia, circadian rhythm disturbances, sleep-related breathing disorders (SRBD), and rapid-eye-movement (REM) sleep behaviour disorder (RBD) are frequent in patients with NDDs. On the other side, poor sleep quality^4, 5^, excessive daytime sleepiness^6^, increased sleep fragmentation^7, 8^, and persistent short sleep duration^5, 9^ predate NDDs diagnosis by one to several years. However, much of the evidence on the association between sleep changes and incident NDDs comes from self-reports^4–6, 9^ and actigraphy data^7–9^ (estimating sleep from rest/activity cycles), yet lacks objective measures of sleep architecture.^10^

The gold standard to assess sleep is polysomnography (PSG)^11^. Scant PSG studies investigated sleep changes and subsequent neurodegeneration^12–15^, mostly focusing on short-term changes in cognitive function but not on long-term NDDs development^14^. In a subset of 321 Framingham Heart Study participants an association between lower REM-sleep percentage and longer REM-sleep latency on the one side, and incident dementia after 12 years mean follow-up on the other side, was identified ^15^. However, home-based PSGs (level II) and not laboratory, personnel-attended PSGs (level I) were used. Additionally, considering solely dementia as outcome may miss some early sleep architecture alterations likely present across the whole NDDs spectrum.

Aggregation of amyloid-β (Aβ) and alpha-synuclein in the brain is a cardinal early step in AD and PD development, respectively. Cross-sectional studies showed cerebrospinal fluid (CSF) total and phosphorilated tau, Aβ and α−synuclein kinetics to be associated with sleep impairment^16, 17^. Moreover, both total sleep deprivation and targeted slow-wave-sleep disruption increased overnight CSF Aβ content^16, 18, 19^. There is also evidence of higher brain interstitial waste product clearance during sleep, maybe due to an increased glymphatic influx^20, 21^. Therefore, the interaction between sleep and kinetics of pathologic proteins’ accumulation/clearance in the brain may provide a common pathogenetic mechanism beyond different pathways leading to neurodegeneration^22^.

Long-term data supporting the relationship between objective sleep changes and future NDDs development are still lacking. Aim of this cohort study was to investigate whether PSG-based sleep features are associated with long-term (i.e., at least five years after baseline) incident NDDs.

## Materials and methods

### Inclusion and exclusion criteria

All patients who underwent PSG at the Sleep Disorders Unit, Department of Neurology, Medical University Innsbruck, Austria, between January 2004 and December 2007 were screened for eligibility. Inclusion criteria were age ≥18 years at the time of PSG (baseline) and ≥five years clinical follow-up. Diagnosis of NDD or established prodromal NDDs, i.e., mild cognitive impairment (MCI) or isolated RBD (iRBD), at baseline or within the first five follow-up years represented an exclusion criterion. The study flowchart is shown in Fig.1.

**Figure 1.**
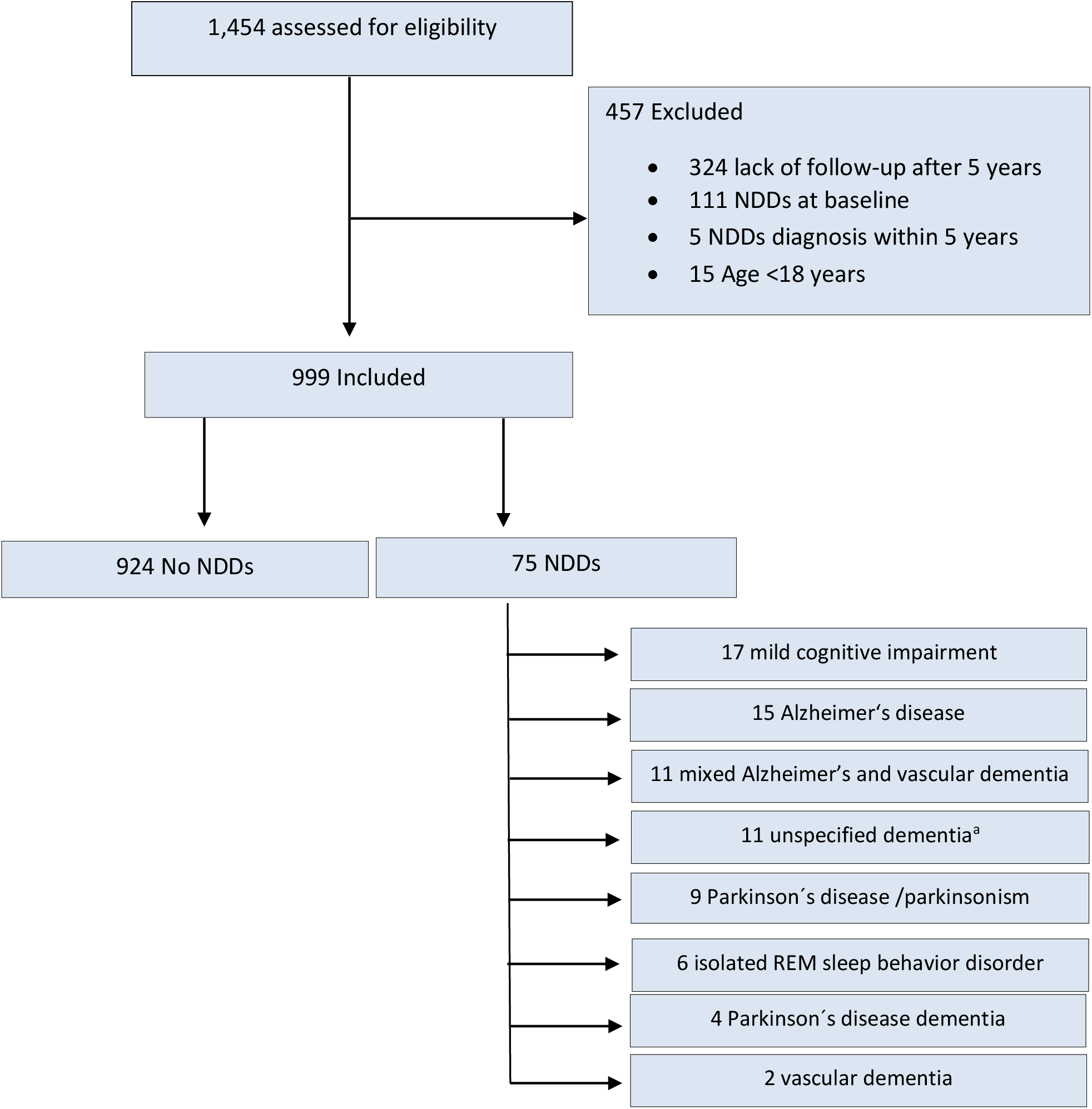
Flowchart of the tudy. Legend: Abbreviations: NDDs: neurodegenerative diseases. ^a^The group unspecified dementia includes patients who received a dementia diagnosis with no available aetiological information.

This study has been conducted in accordance with the declaration of Helsinki, was approved by the local ethics committee and adheres to the STROBE guidelines for observational studies.

### Demographic and clinical data

Demographic and clinical data were retrieved from the hospital’s electronic health records. Demographic data included age, sex, and body mass index (BMI). At baseline, a detailed sleep and medical history were collected, and a full neurological examination was performed. Clinical data included medications and co-morbidities at baseline and over the follow-up time. Sleep disorders were classified according to current version of the International Classification of Sleep Disorders at the time of diagnosis^23, 24^.

### Polysomnography

Eight-hour PSG included: electroencephalography (EEG) with C3, C4, O1, O2, A1, and A2 electrodes; vertical and horizontal electrooculography; electromyography of submental, mental, and both tibialis anterior muscles; single-lead electrocardiography; respiratory monitoring (nasal airflow [thermocouple], thoracic and abdominal piezo-strain gauge belts), tracheal microphone, and transcutaneous oxygen saturation.

Participants underwent PSG for one to three consecutive nights. The first night was used when only one night was available or when the second night was a positive airway pressure (PAP) titration night. In all other cases, the second night was used for analysis.

Sleep stages were manually scored according to Rechtschaffen and Kales. To relate sleep stages to the current AASM standards, S1, S2, and S3+S4 sleep stages are named in this manuscript N1, N2, and N3 sleep, respectively^11^, and movement time is considered as wake. Sleep features of interest in the study included sleep efficiency, sleep latency, REM-sleep latency, percentages of the different sleep stages and wake percentage within sleep period time (SPT, i.e., the time from sleep onset to final awakening), referred to as wake from now on, to improve readability. Respiratory measures included the apnea-hypopnea index (AHI) and oxygen desaturation index (ODI)≥4%. Apneas and hypopneas were scored according to the current AASM definition. Periodic leg movements during sleep indices were calculated using a validated software^25^.

### Outcome measure

The outcome measure was a NDD or established prodromal NDD diagnosis by December 2021. Hospital’s electronic health records of all patients were screened for NDDs in cooperation with regional neurologic and psychiatric memory clinics. NDDs were diagnosed based on clinical and neuropsychodiagnostic assessment, and supportive diagnostic modalities. The following NDDs were identified: AD^26^, mixed Alzheimer’s and vascular dementia^27^, vascular dementia, unspecified dementias, PD, parkinsonism, PD dementia (PDD), iRBD, and MCI^28^.

### Statistics

Statistical analyses were performed using SPSS-26.0 (SPSS, Inc., Chicago-IL, USA) and R-4.1.2. As data were not normal distributed (Shapiro-Wilk test), demographic and clinical data are provided as median (interquartile range, IQR). Mann-Whitney-U (continuous variables), Pearson chi-square, and Fisher’s exact tests (categorical variables) were used to compare baseline characteristics between those who developed NDDs and those who did not. Baseline clinical characteristics were adjusted for age, sex and BMI, and significance was compared with a logistic regression model. P-values<0.05 were considered statistically significant. NDDs risk was estimated using Cox-regression models and Kaplan-Meier analyses. Cox-regression models were used to calculate hazard ratios (HRs) and 95% confidence interval (CI) for each PSG feature. An unadjusted and two adjusted models were built. In adjusted Model 1, HRs were empirically adjusted for relevant clinical variables (age, sex, BMI, and AHI). In Model 2, while maintaining the variables from model 1, clinical covariates (sleep disorders, night of PSG, co-morbidities, medications) were added in a stepwise-regression, and a final model was derived from the selected variables. Data were right censored at the last disease-free follow-up, date of death, or December 1^st^ 2021, whichever came first. The proportional hazards assumption was checked by plotting the Schoenfeld residuals. Since date of NDDs diagnosis does not reflect the true time of disease onset, HRs were also calculated using semi-parametric interval censoring models, built without making any assumptions about the family of baseline distributions. Bootstrap samples (set to 1000) were used for inference on the regression parameters and as estimates for the baseline distribution.

For the Kaplan-Meier analysis, subjects were assigned to quartiles of sleep stages, wake and sleep efficiency. Differences between quartiles were tested pairwise with Log-Rank tests and corrected for multiple comparisons (Bonferroni method). To estimate disease-free survival times, restricted mean survival times were calculated for each quartile.

To rank wake and individual sleep stages’ importance as predictors of incident NDDs and to account for their mutual dependence, permutation feature importance was evaluated with a random survival forest model (RSF), with sleep stages and wake as predictors. The total number of trees was set to 1000, with five features (corresponding to wake and each sleep stage) sampled for splitting and a minimum node size of three.

To investigate whether sleep stages and wake have an additional effect beyond their individual effect, multivariate cox-regression and RSF models were built with all possible combinations. The models’ performances were then evaluated with Harrell’s concordance index.

### Sub-analyses

Six sub-analyses were performed. First, since age is the strongest risk factor for NDDs and sleep architecture changes with aging, HRs were calculated only for subjects ≥50 years at baseline. Secondly, to assess residual confounding on sleep stages from short sleep time, HRs were calculated only for subjects with total sleep time (TST) ≥300 minutes. Thirdly, to account for first night effect, a sub-analysis included only the second-night PSG data. Fourthly, since not all MCI will develop AD, and not all iRBD will phenoconvert to a manifest α-synucleinopathy within an individual’s lifetime, a sensitivity analysis removing these groups was conducted. Additionally, outcomes were stratified into suspected amyloid-pathology (AD, mixed Alzheimer’s and vascular dementia, and amnestic MCI) or suspected α-synucleinopathies (PD, PDD, parkinsonism, and iRBD), and HR were calculated. Vascular dementia, unspecified dementias, unspecified MCI, and non-amnestic MCIs were not included in this sub-analysis. Lastly, since an association between SRBD and NDDs has been suggested, patients with untreated SRBD at baseline who received PAP therapy afterwards (confirmed at a follow-up visit) were compared to those who did not receive PAP therapy.

### Data availability

The datasets generated and analysed during the current study are available from the corresponding authors on reasonable request.

## Results

### Demographics, clinical, and PSG features at baseline

Nine-hundred-ninety-nine patients were included. Seventy-five (7.5%) developed NDDs (Figure 1). Median follow-up time was 12.8 (9.9-14.6) years. Baseline demographic and clinical features of people who developed NDDs vs. those who did not are presented in Table 1. At baseline, those who developed NDDs were significantly older, had more frequently a diagnosis of restless legs syndrome (RLS), hypertension, diabetes mellitus, or depression, and higher rates of antidepressants and dopaminergics use. All subjects treated with dopaminergic agents had RLS. After adjustment for age, sex and BMI, a difference was still present only for depression and antidepressant use.

**Table 1.** Baseline demographic and clinical data of the cohort. <u>Legend:</u> P values < 0.05 are marked in bold. Abbreviations: AHI, apnea-hypopnea index; BMI, body mass index; COPD, chronic obstructive pulmonary disease; IQR, interquartile range; iRBD, isolated REM sleep behavior disorder; NDDs, neurodegenerative diseases; NREM, non-REM sleep; PAP, positive airway pressure; PSG, polysomnography; RBD, rapid eye movement sleep behavior disorder; REM, rapid eye movement; RLS, restless legs syndrome; SRBD, sleep related breathing disorder. ^a^ In case of multiple sleep diagnoses in the same subject, all of them are listed. ^b^PAP treatment at baseline PSG ^c^ The 5 patients with secondary RBD had a diagnosis of narcolepsy type 1. Adjusted odds ratios were adjusted for age, sex, and BMI and adjusted P-vales are calculated from the Walds test.

|  | No NDDs at last follow-up<br>N = 924 | Developed NDDs<br>N = 75 | P value | Adjusted P value | Adjusted odds ratio |
| --- | --- | --- | --- | --- | --- |
| **Demographics** |  |  |  |  |  |
| Age [years], median (IQR) |  |  |  |  |  |
| At baseline PSG | 53.2 (42.6-61.5) | 66.9 (59.3-72.1) | **<0.001** | - | - |
| At last follow-up | 65.7 (55.1-74.2) | 77.1 (71.7-81.7) | **<0.001** | - | - |
| BMI [kg/m<sup>2</sup>] | 27.4 (24.7-30.5) | 28.4 (24.8-32.2) | 0.308 | - | - |
| Men, n (%) | 636 (68.8) | 47 (62.7) | 0.270 | - | - |
| **Sleep diagnosis<sup>a</sup>** |  |  |  |  |  |
| Insomnia, n (%) | 152 (16.5) | 11 (14.7) | 0.688 | 0.463 | 1.31 (0.64-2.69) |
| SRBD, n (%) | 494 (46.5) | 47 (62.7) | 0.124 | 0.850 | 0.95 (0.55-1.63) |
| 5≤AHI≤15 [per hour] | 191 (20.7) | 16 (21.3) | 0.892 | 0.533 | 1.21 (0.66-2.23) |
| 15<AHI≤30 [per hour] | 111 (12.0) | 7 (9.3) | 0.489 | 0.139 | 0.53 (0.23-1.23) |
| AHI>30 [per hour] | 109 (11.8) | 13 (17.3) | 0.159 | 0.955 | 1.02 (0.51-2.05) |
| PAP treated<sup>b</sup> | 83 (9.0) | 11 (14.7) | 0.141 | 0.675 | 1.17 (0.56-2.45) |
| Central disorder of hypersomnolence, n (%) | 31 (3.4) | 1 (1.3) | 0.506 | 0.750 | 0.71 (0.09-5.82) |
| NREM Parasomnia, n (%) | 29 (3.1) | 0 (0) | 0.160 | 0.984 | - |
| REM Parasomnia (excluding iRBD), n (%) | 12 (1.3) | 0 (0) | 1.000 | 0.985 | - |
| Isolated sleep paralysis | 5 (0.5) | 0 (0) | 1.000 | - | - |
| Nightmare disorder | 2 (0.2) | 0 (0) | 1.000 | - | - |
| Secondary RBD<sup>c</sup> | 5 (0.5) | 0 (0) | 1.000 | - | - |
| Circadian rhythm disorders, n (%) | 17 (1.8) | 0 (0) | 0.630 | 0.986 | - |
| RLS, n (%) | 193 (20.9) | 26 (34.7) | **0.006** | 0.126 | 1.53 (0.89-2.65) |
| Sleep related bruxism, n (%) | 29 (3.1) | 0 (0) | 0.160 | 0.983 | - |
| **Isolated symptoms and normal variants, n (%)** |  |  |  |  |  |
| Excessive fragmentary myoclonus | 175 (18.9) | 19 (25.3) | 0.178 | 0.944 | 0.98 (0.54-1.76) |
| Sleep talking | 25 (2.7) | 1 (1.3) | 0.715 | 0.726 | 0.67 (0.07-6.37) |
| **Comorbidities, n (%)** |  |  |  |  |  |
| Hypertension | 360 (39.6) | 41 (54.7) | **0.008** | 0.688 | 0.90 (0.53-1.52) |
| Dyslipidaemia | 298 (32.3) | 28 (37.3) | 0.367 | 0.157 | 0.68 (0.40-1.16) |
| Diabetes mellitus | 100 (10.8) | 14 (18.7) | **0.040** | 0.776 | 1.10 (0.56-2.18) |
| Coronary artery disease | 70 (7.6) | 9 (12.0) | 0.172 | 0.367 | 0.69 (0.31-1.53) |
| Past stroke | 53 (5.7) | 5 (6.7) | 0.740 | 0.235 | 0.54 (0.20-1.49) |
| Epilepsy | 43 (4.7) | 4 (5.3) | 0.789 | 0.455 | 1.55 (0.49-4.86) |
| Renal dysfunction | 10 (1.1) | 2 (2.7) | 0.226 | 0.854 | 1.16 (0.24-5.73) |
| COPD | 82 (8.9) | 10 (13.3) | 0.199 | 0.191 | 1.66 (0.78-3.55) |
| Thyroid disease | 154 (16.7) | 15 (20.0) | 0.459 | 0.919 | 0.97 (0.51-1.84) |
| Depression | 121 (13.1) | 17 (22.7) | **0.021** | **0.016** | **2.17 (1.16-4.07)** |
| **Medications, n (%)** |  |  |  |  |  |
| Antidepressants | 177 (19.2) | 26 (34.7) | **0.001** | **< 0.001** | **2.87 (1.64-5.04)** |
| Neuroleptics | 29 (3.1) | 1 (1.3) | 0.378 | 0.755 | 0.72 (0.09-5.57) |
| Benzodiazepines | 45 (4.9) | 6 (8.0) | 0.236 | 0.757 | 1.16 (0.44-3.05) |
| Nonbenzodiazepine hypnotics | 31 (3.4) | 4 (5.3) | 0.370 | 0.505 | 1.46 (0.48-4.38) |
| Dopaminergic drugs | 53 (5.7) | 13 (17.3) | **<0.001** | 0.125 | 1.79 (0.85-3.75) |
| Antihypertensive drugs | 261 (28.2) | 29 (38.7) | 0.056 | 0.16 | 0.67 (0.39-1.17) |
| Beta-blockers | 124 (13.4) | 10 (13.3) | 0.983 | 0.168 | 0.60 (0.29-1.24) |
| Statins | 166 (18.0) | 18 (24.0) | 0.195 | 0.406 | 0.78 (0.43-1.40) |
| Thyroid substitution therapy | 139 (15.0) | 12 (16.0) | 0.824 | 0.560 | 0.81 (0.40-1.63) |

Polysomnographic features at baseline in people developing NDDs vs. those who did not are reported in Table 2.

**Table 2.** Polysomnographic features at baseline. <u>Legend:</u> Data are presented as median (interquartile range). P values < 0.05 are marked in bold. Abbreviations: AHI, apnea-hypopnea index; AI, apnea index; HI, hypopnea index; NDDs, neurodegenerative diseases; ODI, oxygen desaturation index; PLMS, periodic leg movements during sleep; REM, rapid eye movement; TIB, time in bed; SPT, sleep period time; TST, total sleep time. ^a^ a total of 6 patients in the no NDDs at follow-up group and 3 patients in the developed NDDs group had no REM sleep. ^b^ Data about oxygen saturation was missing for 13 patients in the disease free at last follow-up group, which was excluded.

| Polysomnographic features | No NDDs at last follow-up<br>N = 924 | Developed NDDs<br>N = 75 | P value |
| --- | --- | --- | --- |
| TIB [min] | 480 (480-485) | 480 (477-484) | 0.221 |
| TST [min] | 404 (365-430) | 365 (322-405) | **<0.001** |
| Sleep efficiency [%] | 84.1 (76.5-89.6) | 75.4 (68.1-83.4) | **<0.001** |
| Sleep latency [min] | 14.5 (8.0-27.0) | 17.5 (7.7-31.7) | 0.235 |
| REM latency<sup>a</sup> [min] | 75.0 (56.5-116.4) | 88.5 (63.3-139.1) | **0.042** |
| N1 sleep |  |  |  |
| Time [min] | 61.8 (44.9-86.9) | 69.4 (50.1-111.1) | **0.019** |
| % SPT | 13.6 (9.9-18.9) | 15.7 (11.6-25.9) | **0.002** |
| N2 sleep |  |  |  |
| Time [min] | 216.8 (178.3-247.9) | 193.8 (150.5-233.5) | **0.011** |
| %SPT | 47.6 (39.9-53.7) | 44.4 (34.6-52.4) | **0.042** |
| N3 sleep |  |  |  |
| Time [min] | 11.9 (0-26.7) | 0.1 (0-11.2) | **<0.001** |
| %SPT | 4.30 (0-10.3) | 0.1 (0-3.9) | **<0.001** |
| REM sleep |  |  |  |
| Time [min] | 80.0 (60.3-99.2) | 63.8 (38.5-81.7) | **<0.001** |
| %SPT | 17.5 (13.4-21.6) | 14.1 (8.8-17.6) | **<0.001** |
| Wake |  |  |  |
| Time in SPT [min] | 50.5 (27.5-80.7) | 77.9 (47.9-115.6) | **<0.001** |
| % in SPT | 11.1 (6.1-17.6) | 17.5 (11.4-24.8) | **<0.001** |
| AHI | 3.9 (1.1-14.8) | 5.5 (1.0-19.8) | 0.392 |
| AI | 0.1 (0-1.8) | 0.4 (0-3.1) | **0.015** |
| HI | 3.5 (1.0-10.1) | 4.7 (0.9-10.8) | 0.536 |
| ODI | 1.6 (0.2-7.7) | 2.3 (0.4-11.8) | 0.143 |
| Oxygen levels<sup>b</sup> |  |  |  |
| Mean oxygen saturation [%] | 93 (92-95) | 93 (92-94) | 0.143 |
| Minimal oxygen saturation [%] | 85 (80-88) | 84 (78-87) | 0.051 |
| PLMS index | 8.8 (2.5-25.7) | 26.6 (4.9-48.6) | **<0.001** |

### Polysomnographic features and neurodegeneration risk

After adjusting for covariates, lower sleep efficiency, N3- and REM-sleep percentages were associated with increased NDDs risk (Table 3). Each one-percent decrease in either sleep efficiency, N3- or REM-sleep was associated with a 1.9%, 6.5% or 5.2% increase in NDDs risk, respectively. A 1%-decrease in wake represented a 2.2% reduced risk of incident NDDs. These associations persisted when using a semi-parametric interval-censored model (Table A.1).

**Table 3.** Hazard ratios for the development of neurodegenerative disease using polysomnographic features as continuous variables (one unit decrease). <u>Legend:</u> Data are shown as hazard ratio (95% confidence interval). P values < 0.05 are marked in bold. Abbreviations: AHI, apnea-hypopnea index; ODI, oxygen desaturation index; REM, rapid eye movement sleep; SPT, sleep period time. ^a^Model 1: adjusting for age, sex, body mass index, and AHI. ^b^Model 2: adjusting for age, sex, body mass index, AHI, periodic leg movements during sleep index, and antidepressants. ^c^ Hazard ratio reported for AHI and ODI in model 1 and model 2 are adjusted for only age, sex and body mass index

| **Sleep feature** | **Unadjusted** | **P value** | **Model 1<sup>a</sup>** | **P value** | **Model 2<sup>b</sup>** | **P value** |
| --- | --- | --- | --- | --- | --- | --- |
| Sleep efficiency | **1.044 (1.029-1.055)** | **<0.001** | **1.023 (1.006- 1.039)** | **0.007** | **1.019 (1.002-1.035)** | **0.026** |
| Sleep latency | 0.999 (0.983-1.001) | 0.096 | 0.993 (0.983-1.002) | 0.147 | 1.000 (0.990-1.010) | 0.889 |
| REM latency | 0.998 (0.994-1.001) | 0.290 | 0.999 (0.995-1.002) | 0.549 | 1.000 (0.997-1.004) | 0.614 |
| N1 % in SPT | **0.974 (0.956-0.991)** | **0.002** | **0.964 (0.934-0.990)** | **0.007** | 0.975 (0.944-1.003) | 0.088 |
| N2 % in SPT | **1.024 (1.005-1.042)** | **0.012** | 0.983 (0.961-1.005) | 0.315 | 1.007 (0.983-1.030) | 0.538 |
| N3 % in SPT | **1.127 (1.062-1.160)** | **<0.001** | **1.077 (1.017-1.123)** | **0.010** | **1.065 (1.007-1.118)** | **0.026** |
| REM % in SPT | **1.092 (1.052-1.116)** | **<0.001** | **1.059 (1.018-1.092)** | **0.003** | **1.052 (1.012-1.085)** | **0.009** |
| Wake % in SPT | **0.954 (0.935-0.968)** | **<0.001** | **0.978 (0.958-0.997)** | **0.026** | **0.978 (0.958-0.997)** | **0.026** |
| AHI<sup>c</sup> | 0.995 (1.005-0.984) | 0.386 | 1.003 (1.015-0.990) | 0.616 | 1.002 (0.989-1.014) | 0.757 |
| ODI<sup>c</sup> | 0.998 (1.004-0.992) | 0.675 | 1.000 (1.013-0.986) | 0.989 | 0.999 (1.029-0.985) | 0.940 |

Kaplan-Meier plots are presented in Figure 2. Restricted mean disease-free survival time for the different quartiles of sleep efficiency, single sleep stages and wake are provided in Table A.2. The shortest disease-free survival time was 14.9 (CI 14.6-15.3) years, in people in the lowest N3-sleep quartile and in the highest wake quartile. Patients in the highest wake quartile (>18.6%) had overall the highest NDDs risk compared to other quartiles (78.5%, adjusted HR 3.6 CI 1.2-10.6).

**Figure 2.**
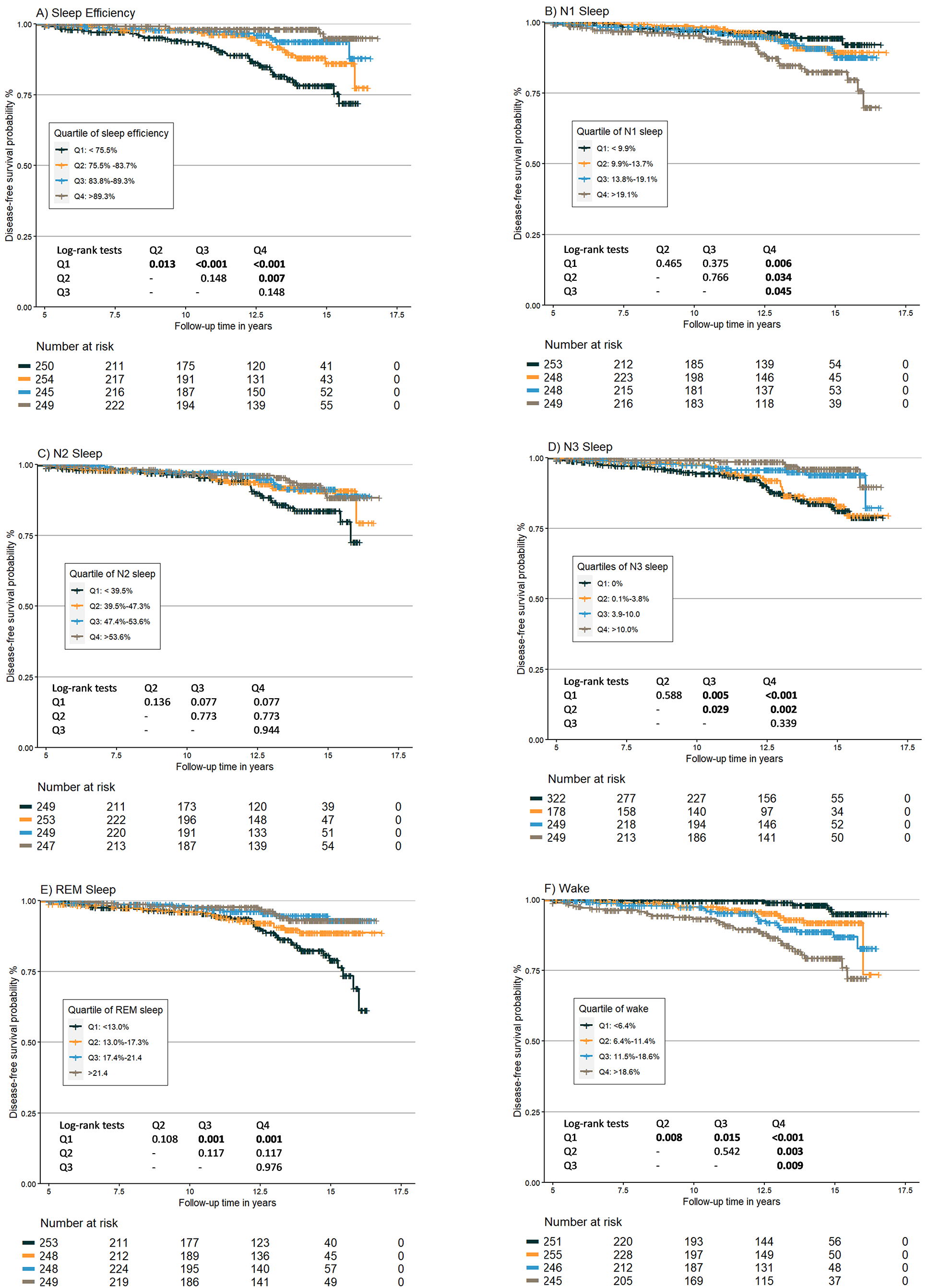
Kaplan-Meier plots by quartiles of polysomnographic features. Legend: Sleep features are expressed as percentages. P-values are corrected for multiple comparisons with the Bonforroni method. P values < 0.05 are marked in bold. Abbreviations: Q, quartile; REM, rapid eye movement sleep.

### Sleep architecture analysis

Multivariate models showed an increased effect of concordance model score when more than one sleep feature was added (C-index 0.58 up to 0.72 for Cox-regression, 0.47 up to 0.72 for RSF, Figure S.1). The best performance was achieved in the Cox-regression model including all sleep stages and wake, and for the combination of N3- and REM-sleep with RSF. The RSF model identified wake as the most important predictor of NDDs, followed by N3- and REM-sleep (Figure S.2).

In the NDDs group, TST at baseline PSG was significantly shorter, whereas by history significantly longer habitual sleep time was reported (Table S.3).

### Sub-analyses

To account for the effect of age, sleep duration and first-night effect, sub-analyses of patients ≥50 years (N=613) sleeping ≥5 hours (N=916), or with second-night PSG study (N=789) yielded consistent results with findings in the whole cohort (Table S.4, Table S.5). When not considering MCI as outcome, only sleep efficiency and wake remained significant (Table A.5, data not shown when not considering as outcome iRBD and both MCI and iRBD). When trying to disentangle risk of different NDDs, reduced sleep efficiency and increased wake were associated with incident suspected amyloid pathology, but no sleep feature was associated with suspected α-synucleinopathies (Table S.6). There was no significant difference in disease-free survival between SRBD patients who received PAP therapy and those who did not (Figure S.3).

## Discussion

This cohort study of 999 patients investigated the association between PSG-measured sleep features and NDDs incidence. Increased wake, reduced sleep efficiency, N3- or REM-sleep percentages were associated with higher long-term risk of incident neurodegeneration. Features’ combination improved NDDs’ risk discrimination compared to single features alone. This is in line with evidence from the literature suggesting that increased wake, as well as reduced REM- and slow-wave-sleep contribute to cognitive decline in older adults^15, 29^.

### Clinical features and neurodegeneration risk

In our cohort, depression prevalence was higher, and antidepressant use more frequent, in those developing NDDs. A previous study found depression late in life to be associated with incident dementia^30^, and depressive symptoms often present early in the course of NDDs. The link between antidepressants intake and incident NDDs is unclear, with contrasting literature findings^10, 31^. Future specifically designed prospective studies should further investigate the relationship between depression/antidepressants’ use and NDDs.

### Polysomnographic features and neurodegeneration risk

#### Sleep efficiency and wake

Our findings extend previous work showing that reduced sleep efficiency is a valuable long-term biomarker for incident NDDs and provide insights into the temporal window during which these changes are detectable. The higher NDDs risk with reduced sleep efficiency in our cohort is in line with previous studies reporting reduced sleep efficiency to be associated with higher amyloid deposition in both cognitively normal individuals^32^, and with incident MCI in older women^33^.

Increased wake was associated with incident NDDs in our cohort. An increase in wakefulness was previously linked to incident dementia and PD^7, 8^. The Framingham Heart Study reported higher dementia risk with longer and more frequent awakenings in subjects with high inflammation level^34^. Increased night wakefulness is indicative of fragmented sleep, which leads to higher oxidative stress and impaired metabolites clearance^20^, potentially contributing to NDDs pathogenesis.

We found a discrepancy between self-reported and PSG-measured sleep duration in those who developed NDDs, with shorter objective TST and longer reported sleep duration. Previous observational and Mendelian randomized studies showed an association between prolonged self-reported sleep duration and incident dementia^35, 36^. These and our findings emphasize the need for objective sleep efficiency and duration measures.

#### REM sleep

In our cohort, reduced REM-sleep was associated to NDDs development after 12.8 years median follow-up time. REM-sleep is likely relevant for sleep-related brain plasticity and emotional memory processing^37, 38^. A reduction in REM-sleep percentage was associated with incident PD^39^. In people with AD, decreased cognitive scores and decreased Aβ-42 CSF levels correlated with decreased REM-sleep^40^. Moreover, a positive association between REM-sleep duration and working memory and executive function was reported^41^. Accordingly, reduced REM-sleep was associated with cognitive deterioration and dementia incidence^14, 15^. Of note, despite different methodologies, our findings are similar to those of the Osteoporotic Fractures in Men Study^14, 42^ concerning what is regarded as low REM-sleep, i.e., ≤13-15%. This cut-off associated with NDDs in our cohort and was previously associated with incident PD^39^ and cognitive decline^14^, suggesting that it may be a valuable NDDs risk marker.

Reduced REM-sleep could represent an early sign of cholinergic system degeneration, which physiologically exhibits greater activity in REM-sleep and is affected by neurodegeneration^43^. In tau-transgenic mice, a decrease in REM-sleep was present before changes in wake or NREM-sleep were observed, and phospho-tau in the sublaterodorsal area negatively correlated with REM-sleep amount^44^. In healthy human adults, overnight changes in brain diffusivity (reflecting changes in water compartmentalization during sleep) correlated with the time spent in REM-sleep^45^, suggesting that REM-sleep duration may affect interstitial clearance.

#### N3 sleep

Although N3-sleep has been implicated in the pathogenesis of NDDs^29, 46^, evidence has been inconsistent^14, 15^. In our study, reduced N3-sleep was associated with incident NDDs. Accordingly, previous cross-sectional studies showed a correlation between reduced N3-sleep and increased CSF total-tau protein in AD patients^40^ and Aβ42 CSF levels in cognitively normal elderly^47^. Moreover, reduced N3-sleep forecasted higher Aβ accumulation over 3.7 years mean follow-up^46^. Adding to that, a retrospective analysis demonstrated higher N3-sleep to be associated with slower motor symptoms’ progression in PD^48^. It has been suggested that during N3-sleep CNS intestinal fluid clearance is improved^21^, and recently a genetic link between N3-sleep and astrocytic water channel aquaporin was found^49^. These findings warrant further investigation into N3-sleep’s role in CNS waste products clearance, and thus in NDDs development.

In our study, EEG included occipital and central but not frontal electrodes (international standards at the time of PSGs). Thus, our data might underestimate N3-sleep compared to current AASM recommendations. Moreover, despite similar NDDs incidence in men and women, the referral rate to our sleep center was biased toward men, who tend to have a higher N3-sleep decline with aging. This might explain absence of N3-sleep in many subjects in our cohort.

### Are specific sleep changes associated with risk of amyloid- or α-synuclein-related NDDs?

Changes in REM- and N3-sleep were significant when considering NDDs as a whole, but stratification into suspected Aß or α-synuclein pathology concealed this association in the latter subgroup. This may be due to the relatively small number of people developing suspected α-synucleionopathies, or may suggest that different sleep structure changes occur in the preclinical stage of these pathologies, maybe due to involvement of different CNS regions. For identification of early sleep changes specifically associated with α- synucleionopathies risk, analysis of sleep microstructure and/or additional PSG information beyond EEG data (e.g., muscular activity) may be needed.

### Sleep disordered-breathing

In our cohort, there was no association between baseline AHI or ODI and incident NDDs. This is in contrast to previous studies, which showed higher incident NDDs in SRBD^12^. In line with our findings, a recent randomized Mendalian study found no causal association between SRBD and incident NDDs^50^. Of note, in our cohort PAP theraphy did not affect incident NDDs risk in patients with SRBD at baseline. However, this was not a population study, was not designed to assess the effect of PAP therapy, and adherence data was not available. Therefore, relevance of our data for what concerns this aspect is limited.

### Strength and limitations

This study has several strengths. We assessed a large cohort using full in-hospital PSG (the gold standard to assess sleep) and provided a comprehensive clinical characterization for a long follow-up time. Analyses were adjusted for various confounders including age, sex, BMI, AHI, antidepressant intake, and sleep disorders. Associations remained robust after multiple sensitivity analyses and after accounting for mutual dependence of the sleep stages.

However, there are some limitations. Firstly, the retrospective design precludes conclusions regarding causality, and our findings need to be confirmed in prospective studies. Moreover, our cohort included patients with sleep complaints. To account for this, diagnosed sleep disorders were added to the survival models as potential confounders. Additionally, MCI might have been missed during routine neurological assessment. Moreover, despite being MCI and iRBD recognized prodromal neurodegenerative conditions, not all people with these conditions will eventually develop NDDs. However, a sensitivity analysis excluding both these groups (data not shown) was consistent with the main findings. No information on social status and education level was available, thus a correction for these variables was not possible. Finally, the limited racial and ethnic diversity (mostly Caucasian patients) restricts generalizability of our findings.

## Conclusions

In this PSG cohort study with up to 16.8 years follow-up, a robust association was found between sleep architectural changes at baseline (i.e., reduced sleep efficiency, REM- or N3-sleep, or increased wake) and incident NDDs. Prospective and Mendelian randomized studies including PSG data are needed to clarify whether this relationship is casual. Future in-depth analysis of micro-architecture sleep features and of additional features recorded during PSG may unveil novel predictive markers of neurodegeneration. The results of the present study add new value to PSG features as preclinical neurodegeneration markers, and have potential implications for the development of prevention strategies, as sleep improvement might slow or even stop neurodegenerative processes.

## Supplementary material

Supplementary material is available at *Brain* online.

## Data Availability

All data produced in the present study are available upon reasonable request to the authors

## Abbreviations

AASM: American Academy of Sleep Medicine
AHI: Apnea Hypopnea
Index AUC: Area Under the Curve
AD: Alzheimer’s Disease
BMI: Body Mass Index
CI: Confidence Interval
C-index: Concordance Index
HR: Hazard Ratio
IQR: Interquartile Range
MCI: Mild Cognitive Impairment
NDDs: Neurodegenerative Diseases
ODI: Oxygen Desaturation Index
PD: Parkinson’s Disease
PDD: Parkinson’s Disease Dementia
PAP: Positive Airway Pressure
PSG: Polysomnography
RBD: Rapid Eye Movement Sleep Behaviour Disorder
RSF: Random Survival Forest
RLS: Restless Legs Syndrome
SRBD: Sleep-related Breathing Disorders
SPT: Sleep Period Time
TIB: Time in Bed
TST: Total Sleep Time

## Supplementary figures and tables legend

**Figure S.1.**
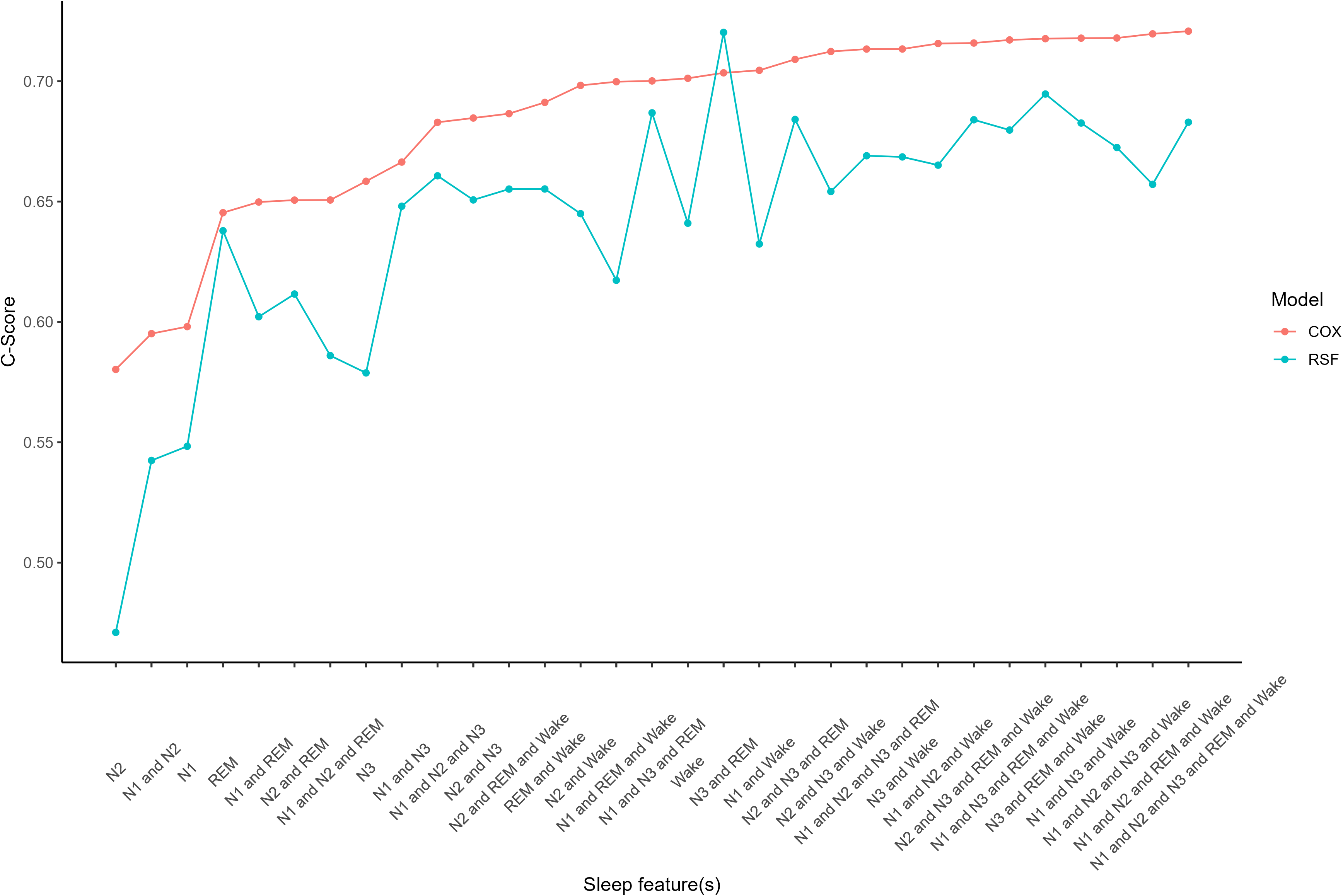
Multivariate analysis of Cox-regression and random survival forest models. <u>Legend:</u> C-Score refers to Harrel’s concordance index Abbreviations: REM, rapid eye movement; COX, Cox regression, RSF random survival forest

**Figure S.2.**
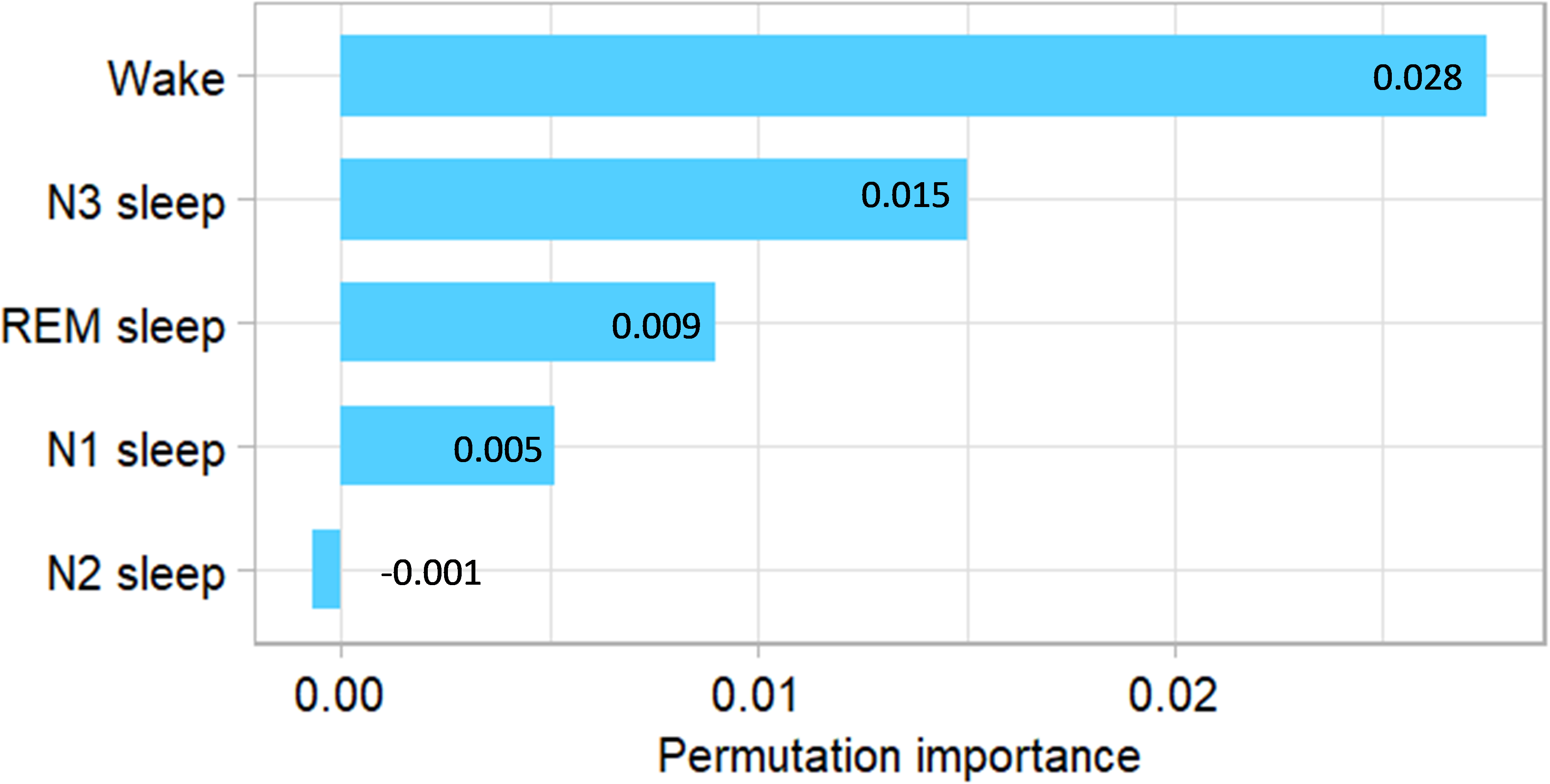
Permutation importance derived from a random forest model. <u>Legend:</u> N2 sleep has a negative value, meaning it negatively impacts the predictions. Abbreviations; REM, rapid eye movement.

**Figure S.3.**
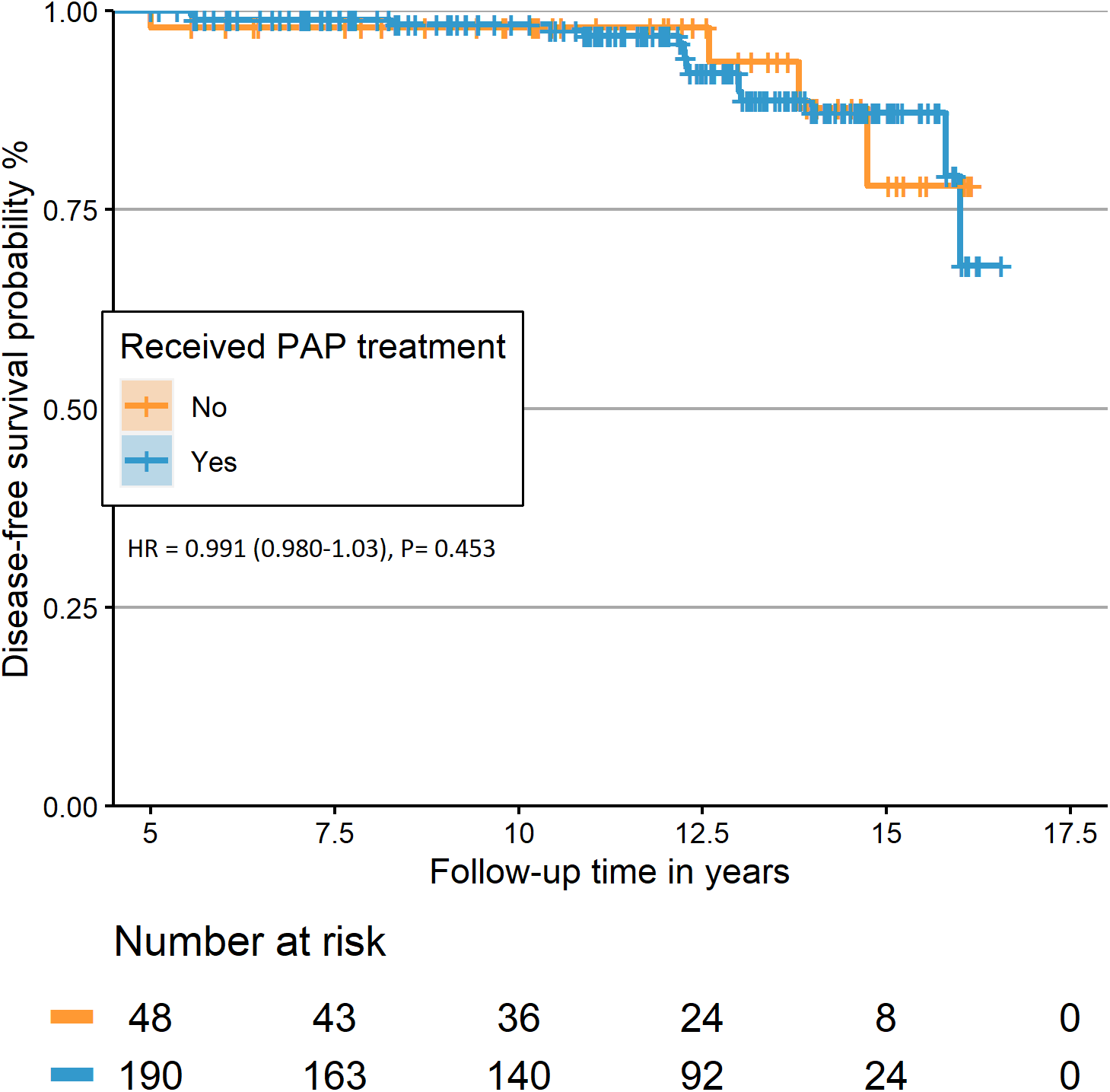
Kaplan-Meier plot of those who received positive airway pressure therapy for sleep-related breathing disorder after the baseline polysomnography. <u>Legend:</u> All the patients received the therapy within 1-2 years post-baseline polysomnography; 15 patients received the treatment after. Hazard ratio was adjusted for age, sex, BMI, antidepressants, and kidney diseases Abbreviations; REM, rapid eye movement, HR: Hazard ratio

**Table S1.** Hazard ratios for development of neurodegenerative disease measured using interval-censoring models. <u>Legend:</u> Data are presented as hazard ratio (standard error). P values < 0.05 are marked in bold. ^a^Model 1: adjusting for age, gender, body mass index, and AHI. ^b^Model 2: adjusting for age, gender, body mass index, AHI, periodic leg movements during sleep index, and antidepressants. Abbreviations: AHI, apnea-hypopnea index; ODI, oxygen desaturation index; REM, Rapid eye movements sleep; SPT, sleep period time.

| **Sleep feature** | **Unadjusted** | **P-Value** | **Model 1<sup>a</sup>** | **P-Value** | **Model 2<sup>b</sup>** | **P-value** |
| --- | --- | --- | --- | --- | --- | --- |
| Sleep efficiency | 0.957 (0.006) | **<0.001** | **0.977 (0.008)** | **0.009** | **1.019 (0.009)** | **0.003** |
| Sleep latency | 1.008 (0.004) | 1.075 | 1.007 (0.005) | 0.218 | 0.999 (0.005) | 0.901 |
| REM latency | 1.002 (0.001) | 0.209 | 1.001 (0.001) | 0.534 | 0.999 (0.001) | 0.614 |
| N1 % in SPT | 1.026 (0.007) | **0.001** | **1.037 (0.151)** | **0.017** | 0.975 (0.245) | 0.113 |
| N2 % in SPT | **0.975 (0.102)** | **0.017** | 0.988 (0.013) | 0.364 | 1.007 (0.128) | 0.566 |
| N3 % in SPT | 0.887 (0.028) | **<0.001** | **0.927 (0.029)** | **0.001** | **1.061 (0.027)** | **0.023** |
| REM % in SPT | 0.915 (0.017) | **<0.001** | **0.943 (0.021)** | **0.007** | **1.050 (0.949)** | **0.011** |
| Wake % in SPT | **1.048 (0.007)** | **<0.001** | **1.022 (0.010)** | **0.032** | **0.978 (0.009)** | **0.024** |
| AHI | 1.005 (1.005) | 0.376 | 0.996 (0.007) | 0.647 | 0.997 (0.008) | 0.897 |
| ODI | 1.001 (0.007) | 0.869 | 1.003 (0.024) | 0.901 | 1.003 (0.020) | 0.885 |

**Table S2.** Restricted mean disease-free survival time for the different quartiles of polysomnographic features. <u>Legend:</u> Significant P values after Bonferroni correction are marked in bold. Abbreviations: CI, confidence interval; Q; quartile; REM, rapid eye movement sleep.

| Sleep feature | Restricted mean disease-free survival time in years (95% CI) | P value, pairwise comparison |  |  | Adjusted increased Risk to develop NDD first (%) |
| --- | --- | --- | --- | --- | --- |
|  |  | Q2 | Q3 | Q4 |  |
| Sleep efficiency |  |  |  |  |  |
| Q1 (< 75.5 %) | 14.9 (14.6-15.3) | **0.040** | **<0.001** | **<0.001** | 65.2 |
| Q2 (75.5%- 83.7%) | 15.8 (15.5- 16.1) | - | 1.000 | **0.007** | 56.8 |
| Q3 (83.8%- 89.3%) | 16.1 (15.8- 16.3) | - | - | 0.957 | 51.8 |
| Q4 (>89.3%) | 16.3 (16.1- 16.5) | - | - | - | Baseline |
| N1 sleep |  |  |  |  |  |
| Q1 (<9.9%) | 16.2 (15.9-16.4) | 1.000 | 1.000 | **0.010** | Baseline |
| Q2 (9.9% -13.7%) | 16.1 (15.8- 16.3) | - | 1.000 | **0.029** | 50.4 |
| Q3 13.8%- 19.1%) | 15.9 (15.6- 16.2) | - | - | 0.154 | 56.6 |
| Q4 (>19.1%) | 15.4 (15.1- 15.8) | - | - | - | 63.2 |
| N2 sleep |  |  |  |  |  |
| Q1 (<39.5%) | 15.2 (14.9- 15.5) | 0.717 | 0.224 | 0.214 | 54.8 |
| Q2 (39.5%- 47.3%) | 15.9 (15.6-16.2) | - | 1.000 | 1.000 | 48.9 |
| Q3 (47.4% - 53.6%) | 15.9 (15.7- 16.2) | - | - | 1.000 | 46.4 |
| Q4 (>53.6%) | 16.1 (15.8-16.3) | - | - | - | Baseline |
| N3 sleep |  |  |  |  |  |
| Q1 (0%) | 14.9 (14.6-15.3) | 1.000 | **0.043** | **<0.001** | 66.6 |
| Q2 (0.1%-3.8%) | 15.8 (15.5-16.0) | - | 0.592 | **0.027** | 68.2 |
| Q3 (3.9-10.0%) | 16.1 (15.8-16.3) | - | - | 1.000 | 55.7 |
| Q4(>10.0%) | 16.3 (16.1-16.5) | - | - | - | Baseline |
| REM sleep |  |  |  |  |  |
| Q1 (< 13.0%) | 15.3 (14.9-15.6) | 0.687 | **0.004** | **0.003** | 61.0 |
| Q2 (13.0% -17.3%) | 15.9 (15.6-16.2) | - | 0.543 | 0.467 | 56.3 |
| Q3 (17.4%- 21.4%) | 16.1 (15.9- 16.4) | - | - | 1.000 | 43.7 |
| Q4 (>21.4%) | 16.2 ( 16.1-16.5) | - | - | - | Baseline |
| Wake |  |  |  |  |  |
| Q1 (<6.4%) | 16.4 (16.3-16.4) | **0.009** | **0.001** | **<0.001** | Baseline |
| Q2 (6.4%- 11.4%) | 15.7 (15.5 -16.0) | - | 1.000 | **0.023** | 72.7 |
| Q3 (11.5%- 18.6%) | 15.8 (15.6-16.1) | - | - | **0.029** | 76.5 |
| Q4 (> 18.6%) | 14.9 (14.6 -15.3) | - | - | - | 78.5 |

**Table S3.** Habitual and PSG sleep time data. <u>Legend:</u> Anamnestic data were available for 782 patients, and missing data were excluded. ^a^The habitual sleep time was calculated by adding the bedtime to the habitual sleep onset time; if not specified, 15 minutes were added to the bedtime. ^b^If the sleep duration was specified by the patient, it was considered otherwise, the difference between the sleep time and wake time was considered the sleep duration. Abbreviations: NDDs: neurodegenerative diseases, PSG: polysomnography

|  | No NDDs at last follow-up | Developed NDDs | P value |
| --- | --- | --- | --- |
| **Habitual sleep Time** |  |  |  |
| Bed time | 22:30:00(22:30:00-22:30:00) | 22:30:00(22:30:00-22:30:00) | - |
| Sleep time<sup>a</sup> | 22:45:00(22:15:00-23:15:00) | 22:45:00(22:15:00-23:18:45) | - |
| Wake time | 06:15:00(05:30:00-07:00:00) | 07:00:00(06:00:00-08:00:00) | - |
| Sleep duration in hours<sup>b</sup> | 7.50(6.5-8.1) | 7.7(7.0-8.7) | **0.033** |
| **PSG sleep time** |  |  |  |
| Light off | 22:31:00(22:16:00-22:45:00) | 22:23:00 (22:15:00 -22:42:30 ) | - |
| Light On | 06:31:00(06:15:00-06:46:00) | 06:25:00 (06:14:30 -06:41:30 ) | - |
| Sleep time | 22:49:00(22:32:30-23:09:00) | 22:47:30 (22:35:15 -23:06:15 ) | - |
| Wake time | 06:08:44(05:48:19-06:29:11) | 06:00:05 (05:34:05 -06:20:09 ) | - |
| Sleep duration in hours<sup>c</sup> | 6.7(6.08-7.1) | 6.1(5.3-6.7) | **<0.001** |

**Table S4.** Hazard ratios Including only people older than 50 years, only people who slept at least 5 hours, or both. <u>Legend:</u> Data are presented as hazard ratios (confidence interval). All models were adjusted for age, gender, body mass index, AHI, periodic leg movements during sleep index, and antidepressants. Significant P values are marked in bold. Abbreviations: AHI, apnea-hypopnea index; ODI, oxygen desaturation; REM, Rapid eye movement sleep; SPT, sleep period time.

| Sleep feature | Patients >50 years | P value | Sleep ≥5 hours | P value | Patients >50 years who slept ≥5 hours | P value |
| --- | --- | --- | --- | --- | --- | --- |
| Sleep efficiency | **1.021 (1.002-1.037)** | **0.022** | **1.030 (1.000-1.058)** | **0.047** | 1.029 (0.997-1.058) | 0.074 |
| Sleep latency | 0.998 (0.988-1.009) | 0.820 | 1.000 (0.986-1.015) | 0.907 | 1.000 (0.985-1.015) | 0.975 |
| REM latency | 1.001 (0.997-1.004) | 0.502 | 1.000 (0.997-1.005) | 0.533 | 1.002 (0.998-1.006) | 0.389 |
| N1 % in SPT | **0.968 (0.937-0.996)** | **0.025** | 0.974 (0.941-1.004) | 0.095 | 0.972 (0.938-1.003) | 0.073 |
| N2 % in SPT | 1.010 (0.985-1.034) | 0.412 | 1.006 (0.979-1.033) | 0.621 | 1.006 (0.978-1.034) | 0.657 |
| N3 % in SPT | **1.076 (1.014-1.124)** | **0.015** | **1.065 (1.005- 1.114)** | **0.033** | **1.07 (1.005-1.122)** | **0.035** |
| REM % in SPT | **1.057 (1.015-1.091)** | **0.006** | **1.048 (1.001-1.088)** | **0.040** | **1.053 (1.004-1.095)** | **0.035** |
| Wake % in SPT | **0.978 (0.957-0.998)** | **0.033** | **0.967 (0.933- 0.997)** | **0.034** | 0.970 (0.936-1.002) | 0.067 |
| AHI | 1.002 (0.989-1.016) | 0.675 | **1.000 (0.986- 1.015)** | **0.890** | 1.003 (0.988-1.018) | 0.670 |
| ODI | 0.980 (0.924-1.031) | 0.453 | 0.998 (0.979- 1.016) | 0.857 | 0.996 (0.936-1.052) | 0.882 |

**Table S5.** Hazard ratios including only people with second night PSG and without taking MCI as an outcome. <u>Legend:</u> Data are presented as hazard ratios (confidence interval). Models^a^ adjusted for age, sex, body mass index, AHI, periodic leg movements during sleep index, antidepressants, and dyslipidemia. Models^b^ adjusted for age, sex, body mass index, AHI, periodic leg movements during sleep index, and antidepressants. P values are marked in bold. Abbreviations: AHI, apnea-hypopnea index; MCI, mild cognitive impairment; ODI, oxygen desaturation; REM, Rapid eye movement sleep; SPT, sleep period time.

| Sleep feature | 2<sup>nd</sup> night PSG<sup>a</sup> | P value | No MCI<sup>b</sup> | P value |
| --- | --- | --- | --- | --- |
| Sleep efficiency | 1.019(0.999-1.037) | 0.062 | **1.022(1.003-1.040)** | **0.026** |
| Sleep latency | 1.000(0.988-1.011) | 0.950 | 0.999(0.988-1.011) | 0.929 |
| REM latency | 1.000(0.996-1.004) | 0.948 | 1.000(0.996-1.004) | 0.962 |
| N1 % in SPT | 0.976(0.941-1.009) | 0.151 | 0.976(0.943-1.007) | 0.135 |
| N2 % in SPT | 1.004(0.977-1.029) | 0.791 | 1.017(0.989-1.043) | 0.234 |
| N3 % in SPT | **1.072(1.009-1.122)** | **0.024** | 1.066(1.000-1.120) | 0.050 |
| REM % in SPT | **1.059(1.014-1.096)** | **0.009** | 1.043(0.998-1.082) | 0.060 |
| Wake % in SPT | **0.978(0.956-0.999)** | **0.041** | **0.976(0.953-0.997)** | **0.024** |
| AHI | 1.003(0.983-1.022) | 0.774 | 1.010(0.993-1.025) | 0.247 |
| ODI | 1.003(0.974-1.031) | 0.850 | 1.008(0.986-1.030) | 0.464 |

**Table S6.** Hazard ratios when the outcome was stratified to suspected α-synucleinopathies or amyloid pathology. <u>Legend:</u> Significant P values are marked in bold. Abbreviations: AHI, apnea-hypopnea index, ODI, oxygen desaturation index: REM, Rapid eye movement a: Adjusting for age, gender, BMI, and AHI, antidepressants, dopaminergic agents, and non-benzodiazepines hypnotics b: Adjusting for age, gender, BMI, AHI, PLMS index, antidepressants, PLMS index, and restless legs syndrome.

| Sleep feature | Alpha synuclinopahties, HR<sup>a</sup> (CI) | P-Value | Amyloid pathology, HR<sup>b</sup> (CI) | P-Value |
| --- | --- | --- | --- | --- |
| Sleep efficiency | 1.003 (0.965-1.040) | 0.846 | **1.031 (1.004-1.056)** | **0.0201** |
| Sleep latency | 1.011 (0.982-1.039) | 0.430 | 0.993 (0.978-1.007) | 0.3338 |
| REM latency | 1.002(0.995-1.009) | 0.507 | 1.001 (0.995-1.007) | 0.5585 |
| N1 % in SPT | 0.956 (0.891-1.013) | 0.135 | 0.999 (0.942-1.052) | 0.9766 |
| N2 % in SPT | 1.002 (0.952-1.050) | 0.913 | 1.029 (0.989-1.066) | 0.1430 |
| N3 % in SPT | 1.060 (0.945-1.157) | 0.302 | 1.060 (0.968-1.138) | 0.1965 |
| REM % in SPT | 1.034 (0.959-1.101) | 0.367 | 1.019 (0.953-1.079) | 0.5606 |
| Wake % in SPT | 0.997 (0.954-1.037) | 0.891 | **0.966 (0.933-0.995)** | **0.0249** |
| AHI | 1.005 (0.976-1.033) | 0.704 | 1.020 (0.995-1.044) | 0.1018 |
| ODI | 0.995 (0.997-1.012) | 0.597 | 1.052 (0.959-1.132) | 0.2668 |

